# Reduction in postoperative opioid requirement associated with use of the NSS-2^®^ Bridge device, a disposable auriculo-nerve field stimulator, and factors affecting the response in cancer patients undergoing abdominal surgical procedures

**DOI:** 10.1101/2022.02.02.22270328

**Authors:** Jacques E. Chelly, Matthew P. Holtzman, David L. Bartlett, Haroon A. Choudry, James F. Pingpank, Amer H. Zureikat, Brittany E. Norton, Senthilkumar K. Sadhasivam, Keith M. Vogt

## Abstract

**Objective:** Determine the effect on opioid use after surgery with use of the NSS-2® Bridge device (NBD^®^) as a field nerve stimulator of the nerves innervating the ear for 5 days.

**Methods:** This was a prospective, randomized, double-blind, placebo-controlled trial investigating the effectiveness of the NBD^®^ in reducing opioid (expressed as oral morphine equivalent; OME, mg) requirement in subjects undergoing abdominal surgery for cancer. A total of 53 subjects randomly assigned to either an active NBD^®^ group or placebo group were included in the analysis. Secondary endpoints included pain using a verbal analogue scale (VAS, 0 = no pain to 10 = worst possible pain), time to ambulation, oral intake, first bowel movement, discharge from the hospital, and tolerability of the NBD^®^. Lastly, functional recovery rated using the 12-item Short Form Survey (SF12) assessed at three months.

**Results:** Use of the NBD® resulted in a 26% overall reduction in OME with no difference in pain level expressed as the area under the curve between postoperative day 1 to 5. respectively). This overall reduction accounts for a 6% reduction in OME in the patients undergoing laparoscopic surgery and a 39% reduction in OME and 25% reduction in pain in patients undergoing open surgery. The tolerability of the device was reported as excellent.

**Conclusions:** Cancer patients have been identified as a population at risk of developing opioid use disorders. This prospective, randomized, double-blind, placebo-controlled study, demonstrated that NBD® may be an effective alternative to the use of opioid postoperatively in patients undergoing abdominal surgery for cancer, especially in especially in patients undergoing open surgery and in elderly.

## INTRODUCTION

Opioid requirement remains a constant concern for patients undergoing abdominal surgery for cancer. Opioid use is often associated with an increased frequency and duration of postoperative ileus [1], length of hospital stay, cost of surgery [2], and more importantly, risk of postoperative opioid use disorder (OUD) [3]. In the past few years, Enhanced Recovery After Surgery (ERAS) protocols have been developed. ERAS protocols include a multimodal approach to perioperative pain management, including the use of regional anesthesia [4-6]. Although such an approach has been proven to reduce pain and opioid use, postoperative pain and opioid requirement remain a concern [7].

The NSS-2 BRIDGE Device (NBD®, Innovative Health Solutions, Versailles, IN, USA) is an auricular, percutaneous, electrical nerve field stimulator. It is small, battery-operated, disposable and stimulates the nerves in the ear for five days (Figure 1). The device is FDA-approved for the treatment of opioid withdrawal symptoms, which include abdominal pain. This characteristic makes the NBD® a potentially interesting alternative for the control of postoperative pain following abdominal surgery [8-10]. Furthermore, a case report suggested that the NBD® may also help reduce postoperative opioid requirement, which in the context of the current opioid crisis, makes the NBD® a non-pharmacological tool for the management of acute pain, a topic of increasing interest [6]. The proposed mechanism of the NBD® is that the device stimulates the terminal branches of the nerves innervating the ear, including the vagal, trigeminal, facial, and glossopharyngeal nerves [11, 12]. Evidence collected in rat studies suggests that stimulation of these nerves indirectly modulates the pain pathways via stimulation of the corresponding cranial nerve nuclei present at the level of the spine and brainstem [13].

**Figure 1:**
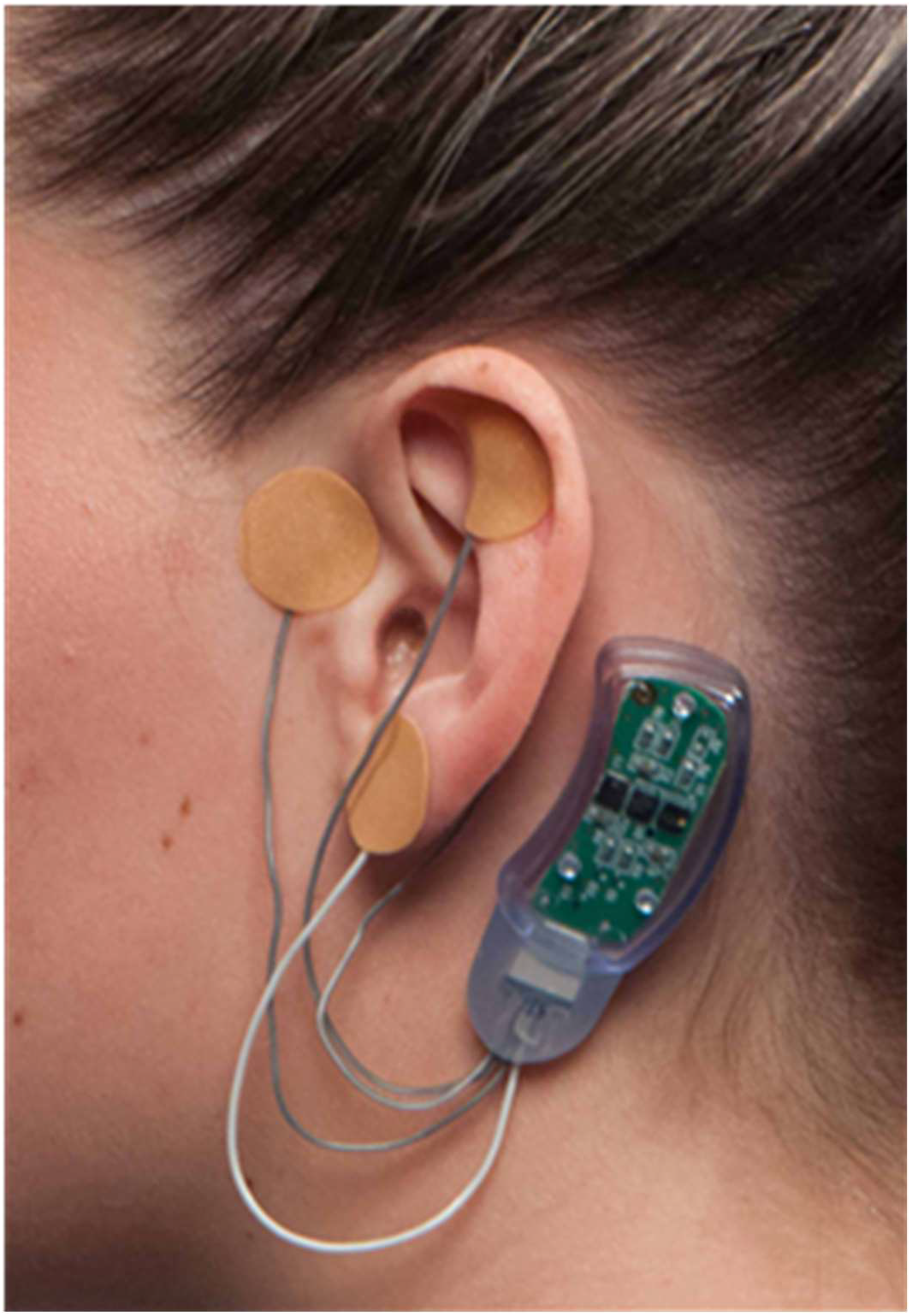
An image of the four affixed and covered electrodes of the NSS-2 BRIDGE®, along with the battery component placed with adhesive behind the ear.

Patients undergoing surgery for cancer have been identified as a population at risk for developing OUD [14]. Since the survival of cancer patients is constantly increasing [15], the ability to minimize exposure to opioids in this patient population using a non-pharmacologic approach to pain may help decrease the risk of OUD following surgery in this population. The current study was designed to assess the potential role of the NBD® in minimizing postoperative opioid requirement in patients with cancer undergoing abdominal surgical procedures and to investigate factors that may impact the effectiveness of the NBD®.

## METHODS

This was a single-center, prospective, randomized, double-blind, placebo-controlled trial conducted at UPMC (University of Pittsburgh Medical Center). This research related to human use has been compliant with all the relevant national regulations, institutional policies, and in accordance with the tenants of the Helsinki Declaration. The protocol was approved by the Institutional Review Board of the University of Pittsburgh and was registered to Clinicaltrials.gov before any eligible patients were approached and asked for consent to participate in the study (STUDY19040260, NCT03555266).

### Recruitment

Subjects were recruited after being scheduled for surgery by a member of the abdominal surgical oncology group at UPMC Shadyside hospital. After obtaining signed informed consent, each subject was randomly assigned to either the active (treatment group) or placebo NBD® group (placebo group). The randomization sequence was determined using computer-generated random numbers.

### Inclusion Criteria

Patients over 18 years of age undergoing elective abdominal surgery performed by a member of the surgical oncology group under general anesthesia were eligible for study inclusion.

### Exclusion Criteria

Patients with a history of active depression, anxiety or catastrophizing, active alcoholism or OUD, or severe chronic pain requiring daily opioids and/or unusual postoperative opioid requirement, defined as a consumption greater than three standard deviations (SDs), were excluded. In addition, the specific exclusion criteria for the NBD® included a history of hemophilia and use of cardiac pacemakers.

### Anesthesia and Perioperative Pain Management

Standard anesthesia care and perioperative pain management included general anesthesia to be induced with intravenous (IV) propofol and rocuronium and maintained with propofol, dexmedetomidine 0.2-0.5 mcg/kg/hr, and ketamine 0.2-1 mg/kg/hr IV infusions, magnesium 2-4g IV, and rocuronium boluses for muscle relaxation to maintain train of four 2/4 twitches. Opioid medications were strictly avoided intraoperatively. The postoperative pain treatment included a peripheral nerve block (quadratus lumborum [16] or paravertebral block [17]) performed prior to surgery, acetaminophen 1g IV or PO every six hours according to each patient’s ability to tolerate oral medication, ketamine 5-10 mg/hr IV infusion for 48 hours, and ketorolac 15 mg IV every eight hours for 48 hours starting in the post-anesthesia care unit. All patients were eligible to receive postoperative opioid medications on an as-needed basis, including oxycodone PO 5 mg every four hours for mild to moderate pain or 10 mg every four hours for severe pain, and hydromorphone IV bolus 0.2-0.5 mg every two hours as needed or patient-controlled administration of hydromorphone 0.2 mg every eight minutes. Ketorolac administration was left at the discretion of the surgeon or acute pain team.

### Treatment Groups

The NBD® was placed upon arrival in the recovery room while the patient was recovering from general anesthesia. According to the subject randomization, an active or placebo NBD® was applied to either the right or left ear by a trained researcher. Except for the research coordinator assigned to placing the NBD®, everyone else was blinded to each subject’s group randomization. The NBD® was left in place for five days or removed early at the subject’s request.

### Data Collection and Outcome Measures

Once each subject gave their informed consent to participate in the study, demographic information and medical history were collected. During the study period, the daily consumption of opioids, acetaminophen, ketorolac, ketamine, oxycodone, or hydromorphone, pain rated using a verbal analogue scale (VAS, 0 = no pain to 10 = worst possible pain), and the number of postoperative nausea and vomiting (PONV) episodes were recorded. Tolerability of the NBD® at the time of placement and one hour after, along with overall patient satisfaction rated on a scale of 0 to 10 (0 = unacceptable, 1-3 = acceptable, 4-7 = good, and 8-10 = excellent) were recorded.

Time to ambulation, time to oral intake, time to first bowel movement, time to discharge from the recovery room, and time to discharge from the hospital were also recorded. Tolerability of the NBD® and patient satisfaction scores were recorded. Lastly, functional recovery scored using the 12-item Short Form Survey (SF12) was assessed at three months.

### Statistics

Data were analyzed using a modified intention to treat analysis [18]. Power was calculated based on a 20-30% loss to follow up of the primary endpoint due to surgery cancellation, postoperative transfer to an intensive care unit, re-intervention because of surgical complications, unusual postoperative opioid consumption beyond three SDs, or undisclosed preoperative opioid use and/or mood disorders that would disqualify the patient from belonging to the same population. Therefore, we estimated that a total of 60 patients would provide 80% power at one side, and a significance level of 0.1 was set. The primary endpoint was total postoperative opioid consumption (OME; mg) over the NBD® five-day stimulation study period or prior to discharge from the hospital. Secondary endpoints included pain (area under the curve over the first five postoperative days); daily pain and opioid consumption; non-opioid consumption and episodes of PONV on postoperative days 1 to 5; time to ambulation, oral intake, and first bowel movement; overall patient satisfaction rated using a 0-10 scale (0 = not satisfied, 10 = most satisfied); and device tolerance rated using a 0-10 scale (0 = unacceptable, 1-3 = acceptable, 4-7 = good, and 8-10 = excellent). At three months, the functional recovery of each subject was assessed using the SF12.

In a pilot study conducted in patients undergoing colorectal surgery, it was suggested that age may affect the effectiveness of NBD® [19]. Therefore, the role of type of surgery (open vs laparoscopic), gender (men vs women), and age (elderly (≥65 years) vs young (<65 years) were also assessed.

Data are reported as mean ± SD. The goal was to assess the effectiveness of the NBD® in reducing postoperative opioid requirements and establish factors that might impact this effectiveness. Therefore, a one-tailed, unpaired t-test was used to compare the NBD® group vs. the placebo group; alpha was set as 0.1.

## RESULTS

A total of 65 subjects provided informed consent. Figure 2 shows a flow chart of all subjects who signed an informed consent. Two patients were excluded due to surgery cancellation, 10 were excluded for inability to properly establish daily postoperative opioid consumption (one subject went back to the operating room for major bleeding, six were admitted to the surgical intensive care unit and received fentanyl for sedation, and two patients required daily opioids outside of three SDs, one in each group), and one subject who requested removal of the device early because it was producing “radiating nerve discomfort,” indicating dislodgment of the device was also excluded. After speaking with the patient, it was unclear when this occurred. Consequently, a total of 53 patients were included in the analysis. Table 1 shows subjects’ demographics and distribution according to the type of surgery (colorectal, pelvic, Whipple, gastric and liver, and ileostomy and hernia, performed either as an open vs. laparoscopic procedure). Table 2 shows the surgery performed on each subject and the approach used (open vs. laparoscopic).

**Figure 2:**
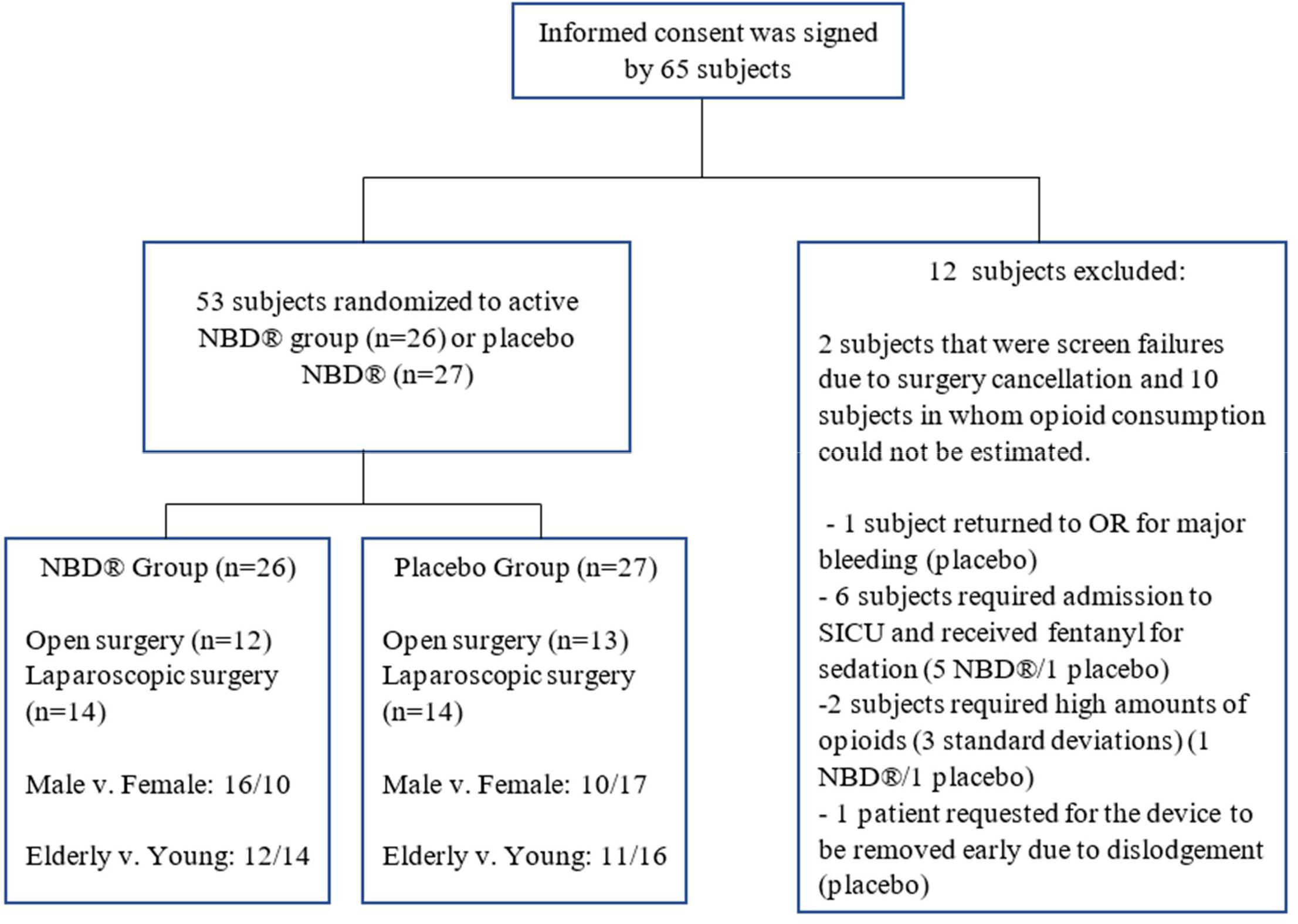
Flow chart of all subjects who consented to participation in the study.

**Table 1.**
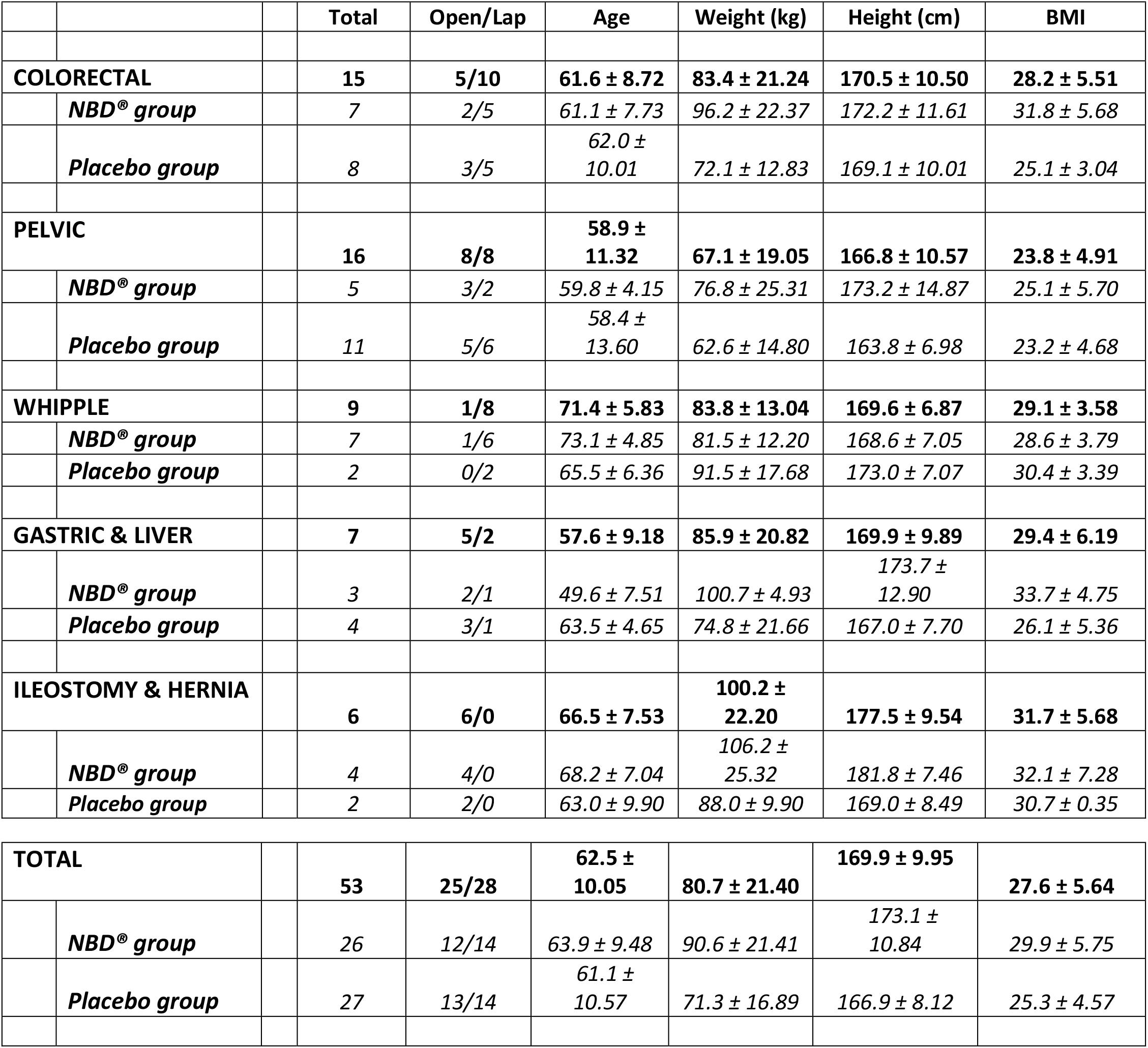
Patients’ demographics and distribution according to the treatment (NBD® vs placebo), the surgery (colorectal, pelvic, Whipple, gastric and liver and ileostomy and hernia) and the type of procedure (open vs laparoscopic). Data expressed as mean ±SD.

**Table 2.**
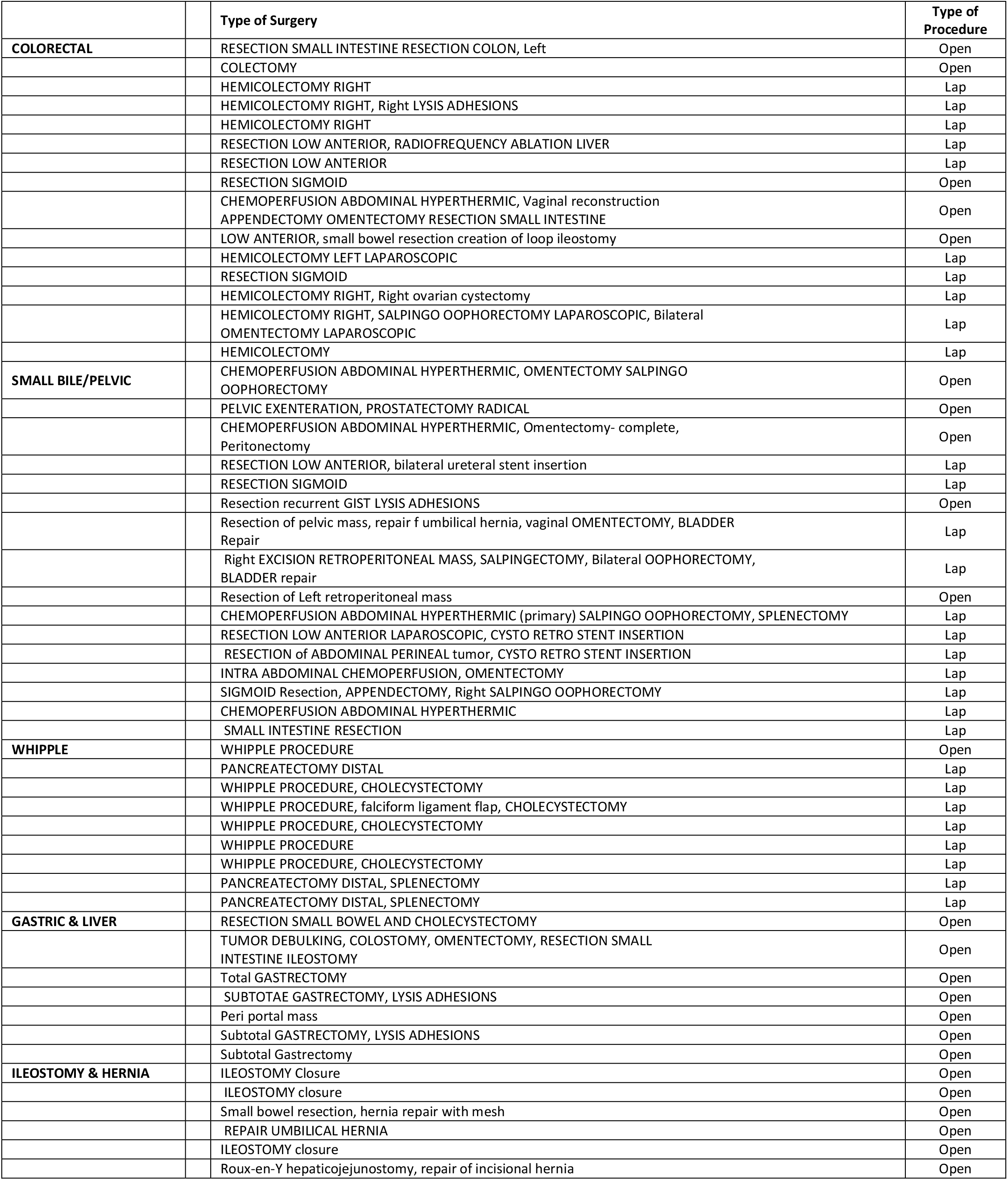
Type of surgery being performed and the type of procedure. Lap = laparoscopic, Open = open.

Table 3 presents the opioid requirement (OME, mg) over the first five days of the recovery period in subjects undergoing both laparoscopic and open surgical procedures and those undergoing open surgical procedures only. Use of the NBD® (n=26) was associated with a non-significant 26% overall reduction of opioid requirement over the five-day study period compared to the placebo group (n=27): 89 mg ± 101 OME mg vs. 120 mg ± 105 OME mg, respectively, (p=0.116). Nineteen percent of subjects in the NBD® group vs. 11% in the placebo group didn’t use any opioids over the five-day study period. The overall reduction in opioid consumption was predominantly attributed to a significant 39% decrease in opioid requirement in subjects undergoing open surgery (n = 25: 91 ± 89 OME in the treatment group (n= 12) vs. 149 ± 112 OME in the placebo group (n= 13), p=0.0854). In contrast, in subjects undergoing laparoscopic procedures (n=28), the use of the NBD® was only associated with a 6% reduction in opioid requirement compared to the placebo group (n=14): 87 mg ± 113 OME vs. 93 mg ± 95 OME, respectively, (p=0.4396). This suggests that the effectiveness of the NBD® was more pronounced in subjects undergoing open surgical vs. laparoscopic procedures.

**Table 3.** Opioids requirement (OME, mg) in subjects undergoing both open and laparoscopic procedures (overall group) and in subjects undergoing only open surgical procedures or only laparoscopic surgical procedures on postoperative day 1, 2, 3, 4 and 5 (1, 2, 3, 4 and 5); OME = oral opioid equivalent expressed in mg.

|  |  | n | 1 | 2 | 3 | 4 | 5 |
| --- | --- | --- | --- | --- | --- | --- | --- |
| **Overall group** |  | 53 |  |  |  |  |  |
|  | NBD® | 26 | 24 ± 27 | 24± 31 | 13 ± 19 | 17 ± 22 | 18 ± 23 |
|  | Placebo | 27 | 38 ± 35 | 27 ± 23 | 22 ± 26 | 23 ± 27 | 23 ± 27 |
|  | p |  | 0.0517* | 0.3326 | 0.0601* | 0.2037 | 0.2937 |
| **Open surgical procedure group** |  | 25 |  |  |  |  |  |
|  | NBD® | 12 | 22 ± 23 | 24± 29 | 15 ± 21 | 18 ± 23 | 15 ± 16 |
|  | Placebo | 13 | 49 ± 41 | 32 ± 25 | 30 ± 28 | 28 ± 29 | 24 ± 32 |
|  | p |  | 0.0295* | 0.2306 | 0.1743 | 0.1897 | 0.2131 |
| **Laparoscopic surgical procedure group** |  | 28 |  |  |  |  |  |
|  | NBD® | 14 | 25 ± 31 | 23 ± 33 | 10 ± 18 | 16 ± 22 | 22 ± 27 |
|  | Placebo | 14 | 28 ± 27 | 22 ± 21 | 15 ± 22 | 18 ± 26 | 21 ± 21 |
|  | p |  | 0.3973 | 0.4509 | 0.2333 | 0.3879 | 0.4677 |

Figure 3 presents pain level by VAS in patients undergoing either laparoscopic or open surgical procedures (n= 53; mean ± SD). Postoperative pain (area under the curve) was similar in both groups (14 ± 7.03 vs. 14 ± 7.66 in the NBD® group vs. placebo group, respectively) and indicates that use of NBD® was associated with a significant reduction in pain on postoperative day 2 (p=0.0917). Figure 4 presents pain scores in the subset of patients undergoing open surgical procedures (n= 25; mean ± SD). The comparison of pain in the NBD® group vs. the placebo group indicated that the use of NBD® was associated with a significant reduction in pain on postoperative days 3 (p=0.0851) and 4 (p=0.0723).

**Figure 3:**
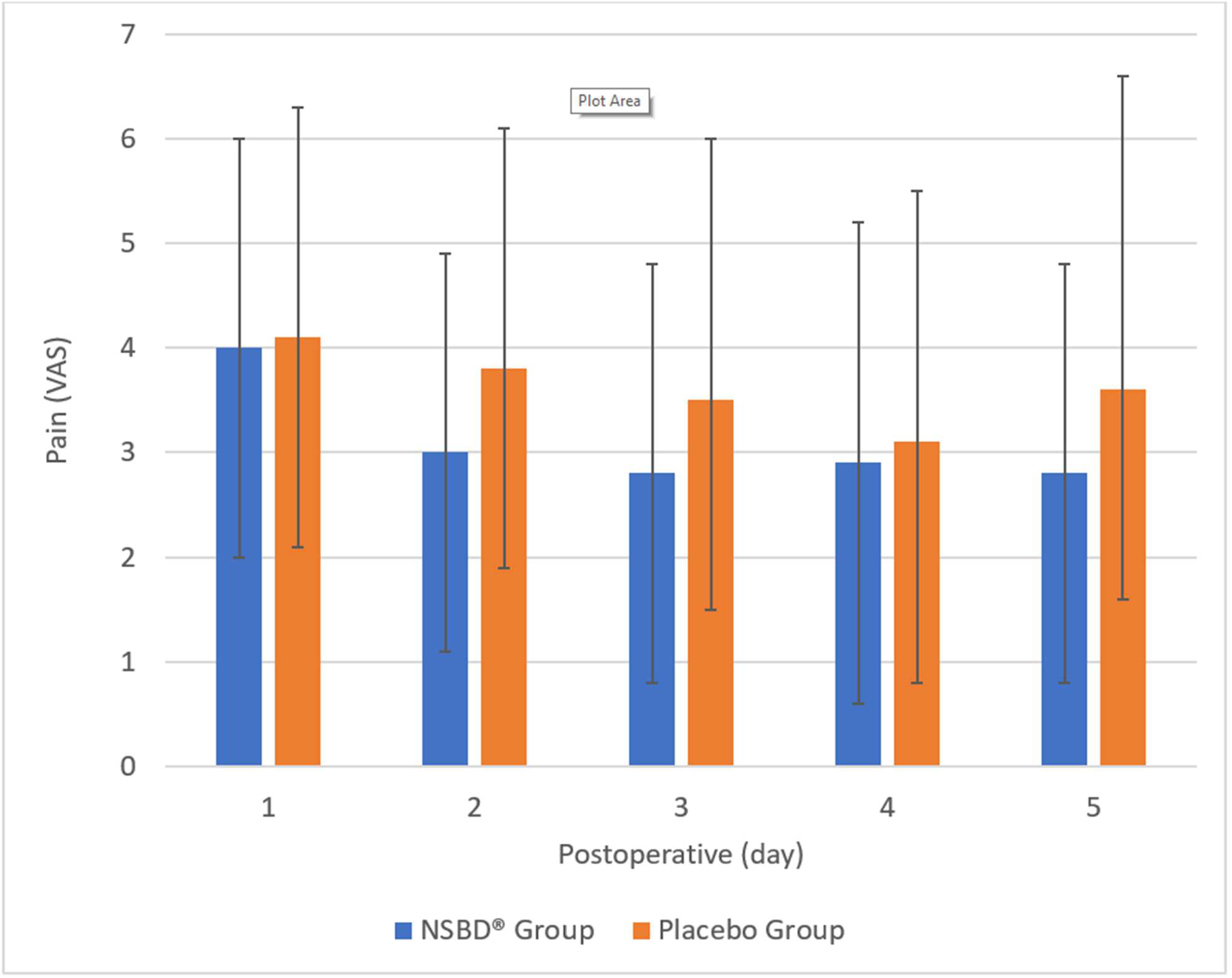
Pain, measured on a 0-10 visual analog scale (VAS), in all patients in the study, having either laparoscopic or open surgical procedures. Values are means, with error bars indicating standard deviation. Postoperative days 1 through 5 are indicated moving left to right.

**Figure 4:**
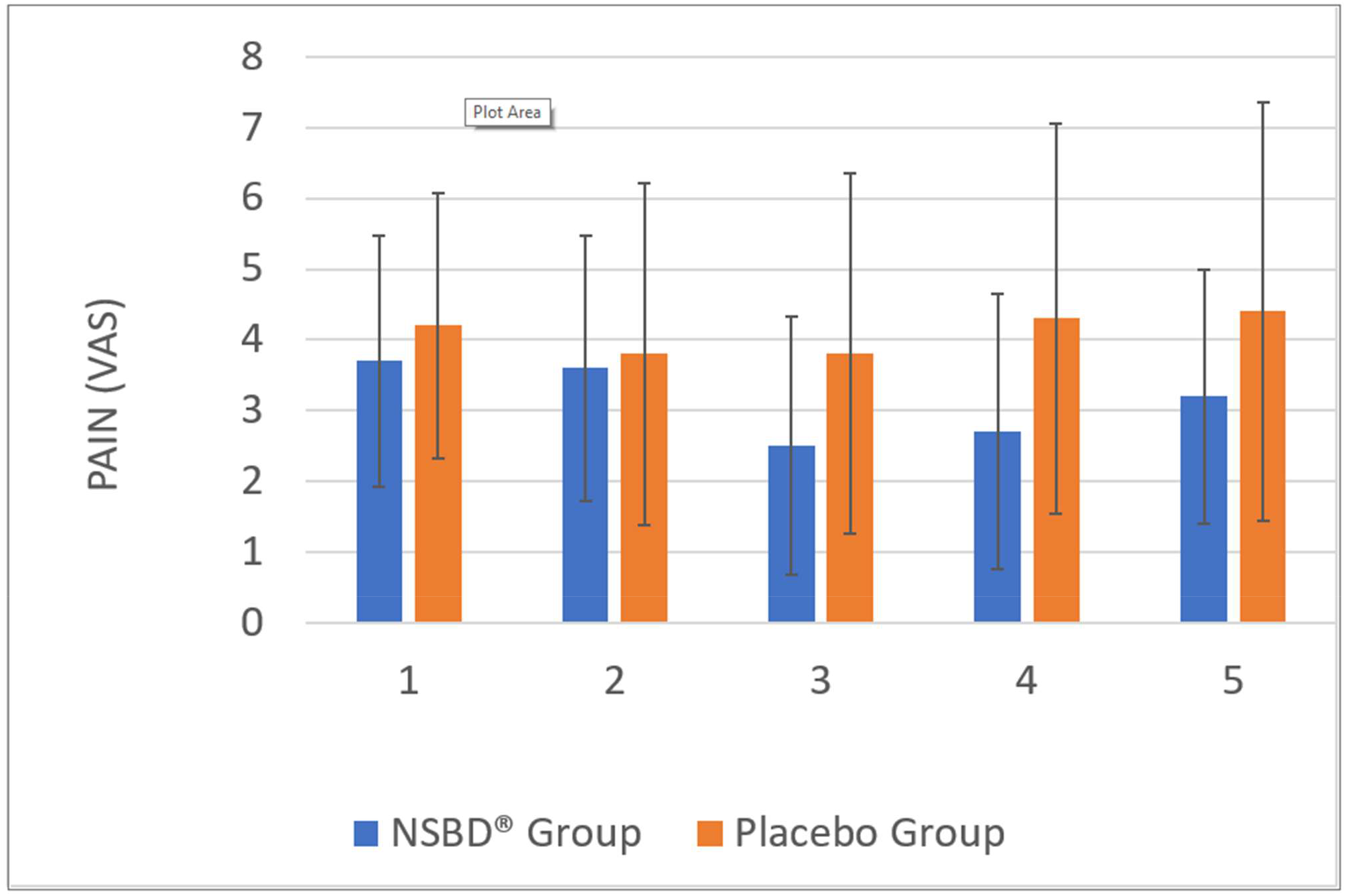
Pain, measured on a 0-10 visual analog scale (VAS), following open abdominal surgery. Values are means, with error bars indicating standard deviation. Postoperative days 1 through 5 are indicated moving left to right.

Except for ketorolac, no differences were recorded in the amount of non-opioid analgesic administered in both the NBD® group and the placebo group. The placebo group received 28% more ketorolac than the NBD® group (p=0.0364).

NBD® effects on overall opioid requirement were similar in males (n=16) vs. females (n=10): 92 ± 112 OME mg vs. 102 ± 85 OME mg, respectively (p=0.4313). NBD® effects on overall opioid requirement was 56% greater in elderly (n=12) compared to younger subjects (n=14) : 57 ± 70 OME mg vs. 116 ± 117 OME mg, respectively (p=0.0709).

Time to discharge from the recovery room was similar in both groups (192.3 ± 48.06 mins for the subjects in the treatment group vs. 207.1 ± 84.38 min in the placebo group, p = 0.2187). Incidence of PONV over the five-day study period between the treatment group and the placebo group was found to be similar (1.08 ± 1.70 episodes vs. 1.52 ± 2.78 episodes, p=0.2451). Time to oral intake and time to ambulation was similar in both groups (45 hrs. ± 41.8 hrs in the treatment group vs. 35 ± 37.4 hrs in the placebo group, p=0.1950 and 26 ± 22.6 hrs in the treatment group vs. 24 ± 18.9 hrs in the placebo group, p=0.4028, respectively). Time to first bowel movement was longer in the treatment group compared to the placebo group (79 ± 32.6 hrs vs. 62 ± 36.1 hrs, p = 0.0755). There was no difference in time to discharge from the hospital between the groups. (144 hrs. ± 48.7 hrs vs. 122 ± 67.5 hrs, respectively).

At the time of device placement, overall device tolerability was reported as excellent overall in 91% of the subjects (86% for the NBD® group vs. 94% in the placebo group). This number increased to 97% one hour after the placement. Two subjects in the NBD® group and two subjects in the placebo group asked for the device to be removed prior to the end of the study period. In the remaining subjects, device tolerability was reported as excellent.

Overall patient satisfaction was very high and similar in both the treatment and placebo groups (8.7 ± 1.49 in the NBD® group vs. 9.0 ± 1.13 in the placebo group (p = 0.2647). Lastly, SF12 scores were found to be similar in both groups (mental 37.5 ± 8.28 vs. 39.9 ± 10.03 and physical 55.2 ± 7.45 vs. 52.1 ± 8.25, in the NBD® group vs. the placebo group, respectively).

## DISCUSSION

Our study suggests that the use of the NBD® may be an effective non-pharmacological technique to reduce postoperative opioid requirements in cancer patients undergoing open abdominal surgery. This is significant in the current opioid crisis and the role of perioperative opioid use in the development of OUD. Although the recorded effects of the NBD® on opioid requirement were only 39% in patients undergoing open surgery compared to placebo, it is important to recognize that these effects were recorded in the presence of a 35% increase in ketorolac consumed in the placebo group. Our protocol didn’t control for the amount of ketorolac received by each patient because its administration was dictated by the surgeon based on postoperative creatinine and associated risk of bleeding. Therefore, the potential benefits of the NBD® are most likely underestimated. Recent case series have reported a greater than 60% reduction in opioid requirement associated with the use of the NBD® [10, 20].

The ear is uniquely innervated by the branches of the trigeminal, facial, glossopharyngeal, and vagal cranial nerves and the superficial cervical plexus due to its embryological origin [21, 22]. The nuclei of these cranial nerves are located in the brainstem and closely connected to a number of structures, including the tractus solitaries and structures involved in the limbic system such as the thalamus and hypothalamus, amygdala, and rostral ventral medulla [13]. The proposed mechanism of action of the NBD® is that the direct auricular percutaneous electrical nerve field stimulation generates action potentials from different areas of the ear that are transmitted on the corresponding cranial nerve nuclei. At the level of the spine and brainstem, these stimulations activate other brain structures and pathways [13]. This concept is supported by a study investigating the role of the NBD® in reducing post-inflammatory hyper-analgesia in rats [23]. In these animal models, using a stereotaxic approach, it was demonstrated that the auricular percutaneous electrical nerve field stimulation induced by the NBD® led to specific electro firing in the central nucleus of the amygdala and spinal cord.

This study also confirms that NBD® tolerability is excellent in most subjects [10, 22]. The NBD® was placed in the recovery room while the patients were still lightly sedated because the goal was to focus on the role that NBD® may play in reducing the use of opioids postoperatively which represents a period when the patient, not the health care provider determine his/her needs for opioids. Although, it would have been possible to place NBD® prior to surgery, doing so would have required the need to account for arbitrary use of opioids during surgery. In awake, non-anesthetized subjects, placement of the NBD® is sometimes associated with a mild impression of “tickling” for a very short duration (1-3 min) immediately after the placement of electrodes and activation of the NBD®. This sensation disappears quickly (Chelly, personal communication).

Our data suggest that the effectiveness of the NBD® is greater in patients undergoing open surgical procedures compared to those undergoing laparoscopic surgical procedures. Blank et al. also reported [19] that the NBD® was not effective in reducing postoperative opioid requirements in patients undergoing laparoscopic colorectal surgery. One possible explanation for the decrease of NBD® effectiveness in patients undergoing laparoscopic procedures may be that laparoscopic procedures involve local pain mechanisms, whereas open surgery involves both peripheral and central mechanisms. Another possible explanation is that laparoscopic procedures are less invasive than open surgery. Accordingly, laparoscopic surgery should be associated with less pain compared to open surgery. However, in the context of a multimodal approach to postoperative pain, we found that the level of pain (expressed as the area under the curve from postoperative day 1 to day 5) was similar (15 ± 8.2 open vs. 12 ± 7.4 laparoscopic, respectively, p=0.1702). Therefore, the pain mechanism involved in both types of procedures may be the predominant factor.

Age was another factor found to impact the effectiveness of the NBD®. Thus, the effectiveness of the NBD® on opioid requirement was higher in elderly patients undergoing open surgical procedures compared to those undergoing laparoscopic surgical procedures. The mechanism of such a difference is yet to be established. A study including a larger number of patients undergoing both open and laparoscopic procedures of various ages is required to confirm the role that age and type of surgical procedure play in impacting the effectiveness of NBD®.

## Limitations

Although the study design was randomized and placebo-controlled, it was conducted in a specific surgical population. Additional data is required to validate the ability of NBD® to reduce postoperative opioid requirements. Furthermore, the study was not powered for assessing the factors modulating the effectiveness of NBD® to reduce postoperative opioid requirements. Additional data obtained in a larger population of patients would be required to validate the ability of NBD® to reduce postoperative opioid requirements and to determine the factors affecting the response.

## CONCLUSION

Our study suggests that the use of the NBD® may represent an effective alternative to control postoperative pain and opioid consumption in patients undergoing abdominal surgical oncology procedures, especially when an open surgical approach is used. However, additional studies are required to confirm these findings.

## Data Availability

All data produced in the present work are contained in the manuscript

## AUTHOR CONTRIBUTIONS

Jacques E. Chelly, MD, PhD, MBA: Conception and design, data acquisition, data analysis and interpretation, statistical analyses, drafting of the manuscript, critical revision of the manuscript, and supervision.

Matthew P. Holtzman, MD: Drafting of the manuscript, critical revision of the manuscript, and supervision.

David L. Bartlett, MD: Conception and design, drafting of the manuscript, critical revision of the manuscript and supervision.

Haroon A. Choudry, MD: Drafting of the manuscript, critical revision of the manuscript, and supervision.

James F. Pingpank, MD: Drafting of the manuscript, critical revision of the manuscript, and supervision.

Amer H. Zureikat, MD: Drafting of the manuscript, critical revision of the manuscript, and supervision.

Brittany E. Norton: Data acquisition, data analysis and interpretation, statistical analyses, drafting of the manuscript, and critical revision of the manuscript.

Senthilkumar K. Sadhasivam, MD, MPH, MBA: Review of the manuscript and critical revision

Keith M. Vogt, MD, PhD: Review of the manuscript and critical revision

## INFORMED CONSENT

Informed consent was obtained from all individuals included in this study.

## ACKNOWLEDGMENTS

The authors want to acknowledge Mrs. Christine Burr for editing this manuscript.

